# Exploring potential causal genes for uterine leiomyomas: A summary data-based Mendelian randomization and FUMA analysis

**DOI:** 10.1101/2022.03.06.22271955

**Authors:** Yuxin Dai, Xudong Liu, Yining Zhu, Su Mao, Jingyun Yang, Lan Zhu

## Abstract

**Objective:** To explore potential causal genetic variants and genes underlying the pathogenesis of uterine leiomyomas (ULs).

**Methods:** We conducted the summary data-based Mendelian randomization (SMR) analysis and performed functional mapping and annotation using FUMA to examine genetic variants and genes that are potentially involved in the pathogenies of ULs.

Both analyses used summarized data of a recent genome-wide association study (GWAS) on ULs, which has a total sample size of 244,324 (20,406 cases and 223,918 controls). For the SMR analysis, we performed separate analysis using CAGE and GTEx eQTL data.

**Results:** Using the CAGE eQTL data, our SMR analysis identified 13 probes tagging 10 unique genes that were pleiotropically/potentially causally associated with ULs, with the top three probes being ILMN_1675156 (tagging *CDC42*, P_SMR_=8.03×10^−9^), ILMN_1705330 (tagging *CDC42*, P_SMR_=1.02×10^−7^) and ILMN_2343048 (tagging *ABCB9*, P_SMR_=9.37×10^−7^). Using GTEx eQTL data, our SMR analysis did not identify any significant genes after correction for multiple testing. FUMA analysis identified 106 independent SNPs, 24 genomic loci and 137 genes that are potentially involved in the pathogenesis of ULs, seven of which were also identified by the SMR analysis.

**Conclusions:** We identified many genetic variants, genes, and genomic loci that are potentially involved in the pathogenesis of ULs. More studies are needed to explore the exact underlying mechanisms in the etiology of ULs.

## Introduction

Uterine leiomyomas (ULs), also called myomas or uterine fibroids, are benign tumors in the smooth muscle tissue in myometrium[1, 2]. The overall prevalence of UL is about 70% in women of reproductive age, and approximately 25% of UL patients suffer from apparent clinical symptoms and require treatment[3]. ULs are the most prevalent benign tumor in female reproductive tract and the leading indication for hysterectomy. ULs represent a major cause of morbidity in women of childbearing age and account for excessive menstrual bleeding, pelvic pain or pressure, infertility, and pregnancy complications[4]. To date, the only definitive treatment for ULs, including the familial subtype, is hysterectomy, which creates a great challenge if fertility preservation is desired. ULs also cause tremendous economic burden. For example, the annual cost of ULs in the US alone, including direct medical costs and indirect financial losses, is estimated to be up to $34.4 billion, higher than the combined cost of breast and colon cancer[5].

UL is a complex, multi-factorial gynecological benign disease with highly variable tumor size, tumor location and clinical manifestations. Many factors have been reported to be associated with the risk of ULs, including biological, demographic, reproductive and lifestyle factors[6-8]. Furthermore, previous studies also suggested that genetics plays an important role in the pathogenesis of ULs. For example, African-American women, or generally women with African origin, are more predisposed to develop ULs, with a prevalence as high as 80% [9], suggesting ethnicity-specific factors, potentially ethnicity-specific genetic structure, underlying the pathogenesis of ULs. Familial clustering between first-degree relatives and twins was also observed as well as multiple inherited syndromes in which fibroid development occurred[10, 11]. Moreover, many GWAS and candidate gene studies have identified several genetic variants/loci associated with the susceptibility of ULs[12-17]. However, the role of putative risk factors and the underlying biological mechanisms underpinning ULs remain largely unclear, which has contributed to the slow progress in the development of effective treatment options for ULs. More studies are needed to explore genetic variants/genes that are potentially causally associated with ULs to better understand the pathogenesis of ULs.

Mendelian randomization (MR) uses genetic variants as the proxy to randomization. Recently, it has been widely adopted to explore pleiotropic/potentially causal effect of an exposure on the outcome (e.g., ULs)[18]. Confounding and reverse causation, which are commonly encountered in traditional association studies, can be greatly reduced by MR. This method has been successful in identifying gene expression probes or DNA methylation loci that are pleiotropically/potentially causally associated with various phenotypes, such as neuropathologies of Alzheimer’s disease and severity of COVID-19[19, 20].

In this paper, we attempted to prioritize genes that are potentially causally associated with ULs through a summary data-based MR (SMR) approach. We also performed functional mapping and annotation to further explore genetic variants and genomic loci that are potentially involved in the pathogenesis of ULs.

## Methods

### GWAS data for ULs

The GWAS summarized data for ULs were provided by a recent genome-wide association meta-analysis of ULs[14]. The results were based on meta-analyses of ULs using data from four population-based cohorts (Women’s Genome Health Study, UK Biobank, Queensland Institute of Medical Research, and North Finnish Birth Cohort), with a total sample size of 244,324 (20,406 cases and 223,918 controls). Genotyping was done on different platforms, and imputation was performed using the reference panel from the 1000 Genomes Project European dataset (1000G EUR) Phase 3 or the Haplotype Reference Consortium (HRC) panel. For each cohort, logistic regression or linear mixed model association analysis was done, assuming an additive genetic model and adjusting for age, BMI, and/or the first five principal components, and/or array type, as appropriate. The GWAS summarized data can be downloaded at http://ftp.ebi.ac.uk/pub/databases/gwas/summary_statistics/GCST009001-GCST010000/GCST009158/.

### eQTL data

The SMR analysis used cis-eQTL genetic variants as the instrumental variables (IVs) for gene expression. We performed separate SMR analysis using eQTL data from two sources. Specifically, we used the V7 release of the GTEx eQTL summarized data for uterus[21], which included 70 participants, and the CAGE eQTL summarized data for whole blood[22], which included 2,765 participants. The eQTL data can be downloaded at https://cnsgenomics.com/data/SMR/#eQTLsummarydata.

### SMR analysis

The SMR analysis was done using the software SMR. Detailed information regarding the SMR method was reported elsewhere[28]. The existence of linkage in the observed association was assessed using the heterogeneity in dependent instruments (HEIDI) test. P_HEIDI_<0.05 means rejection of the null hypothesis. That is, the observed association could be due to two distinct genetic variants in high linkage disequilibrium with each other. We adopted the default settings in SMR (e.g., P_eQTL_ <5 × 10^−8^ and minor allele frequency [MAF] > 0.01) and used false discovery rate (FDR) to adjust for multiple testing. The SMR analytic process is illustrated in **Figure 1**.

**Figure 1.**
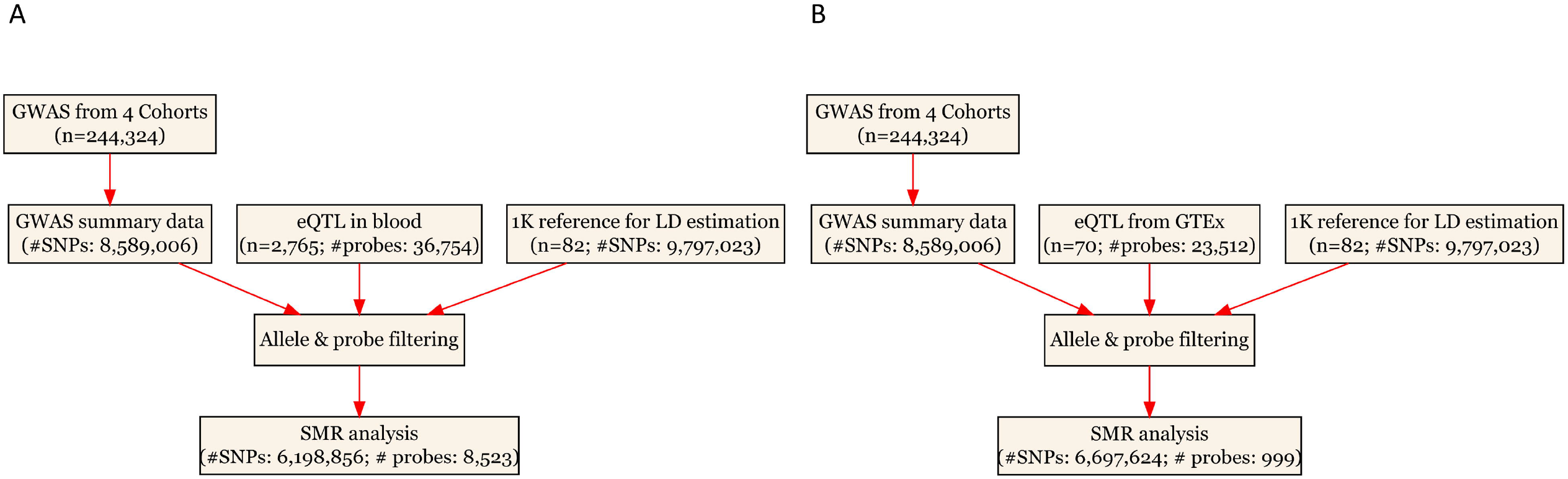
Flow chart for the SMR analysis. A) SMR analysis using CAGE eQTL data from blood; and B) SMR analysis using GTEx eQTL data eQTL, expression quantitative trait loci; GWAS, genome-wide association studies; LD, linkage disequilibrium; SMR, summary data-based Mendelian randomization; SNP, single nucleotide polymorphism

### FUMA analysis

To better understand the genetic mechanisms underlying ULs, we also conducted a FUMA analysis to functionally map and annotate the genetic association, again using the GWAS summary results of ULs. FUMA is an on-line platform that integrates information from multiple resources for easy implementation of post-GWAS analysis, such as functional annotation and gene prioritization[23]. It has two processes, SNP2GENE, which annotate SNPs regarding their biological functions and map them to genes, and GENE2FUNC, which annotates the mapped genes in biological contexts. In SNP2GENE, we performed both positional mapping and eQTL mapping using GTEx v8 of whole blood and uterus. We selected all types of genes in gene prioritization and adopted the default settings otherwise (e.g., maximum P-value of lead SNPs being 5×10^−8^ and r^2^ threshold for independent significant SNPs being 0.6). In GENE2FUNC, we adopted the default settings (e.g., using FDR to correct for multiple testing in the gene-set enrichment analysis).

Data cleaning and statistical/bioinformatical analysis was performed using R version 4.1.2 (https://www.r-project.org/), SMR (https://cnsgenomics.com/software/smr/), and FUMA (https://fuma.ctglab.nl/).

## Results

### Basic information of the summarized data

In the SMR analysis, the CAGE eQTL has a much larger number of participants than that of the GTEx eQTL data (2,765 vs. 70), so is the number of eligible probes (8,523 vs. 999). After checking allele frequencies among the datasets and LD pruning, there were more than 6 million eligible SNPs in each SMR analysis. In the FUMA analysis, about 8.6 million SNPs were used as the input. The detailed information was shown in **Table 1**.

**Table 1.** Basic information of the eQTL and GWAS data.

| **Data Source** | **Total # of participants** | **Number of eligible genetic variants or probes** |
| --- | --- | --- |
| **eQTL data** |  |  |
| **CAGE** | 2,765 | 8,523 |
| **GTE<sub>x</sub>** | 70 | 999 |
| **GWAS data for SMR analysis** |  |  |
| **WGHS** | 3,375/9,465 | - |
| **NFBC** | 363/5,000 | - |
| **QIMR** | 1,484/3,701 | - |
| **UKBB** | 15,184/205,752 | - |
| **Total** | 20,406/223,918 | CAGE: 6,198,856; GTE<sub>x</sub>: 6,697,624 |
| **GWAS data for FUMA analysis** | 20,406/223,918 | 8,589,006 |

GWAS: genome-wide association studies; QTL, quantitative trait loci; WGHS, Women's Genome Health Study; QIMR, Queensland Institute of Medical Research; UKBB, UK Biobank, NFBC, North Finnish Birth Cohort

### Pleiotropic association with UL

Using the CAGE eQTL data, our SMR analysis identified 13 probes tagging 10 unique genes that were pleiotropically/potentially causally associated with ULs, with the top three probes being ILMN_1675156 (tagging *CDC42*, P_SMR_=8.03×10^−9^), ILMN_1705330 (tagging *CDC42*, P_SMR_=1.02×10^−7^) and ILMN_2343048 (tagging ABCB9, P_SMR_=9.37×10^−7^; **Table 2**). There were three probes tagging *CDC42* (**Figure 2**) and two probes tagging *ABCB9* (**Figure 3**) that showed significant pleiotropic association with ULs. Using GTEx eQTL data, we did not identify any genes that were pleiotropically/potentially causally associated with ULs after correction for multiple testing (**Table 2**).

**Table 2.**
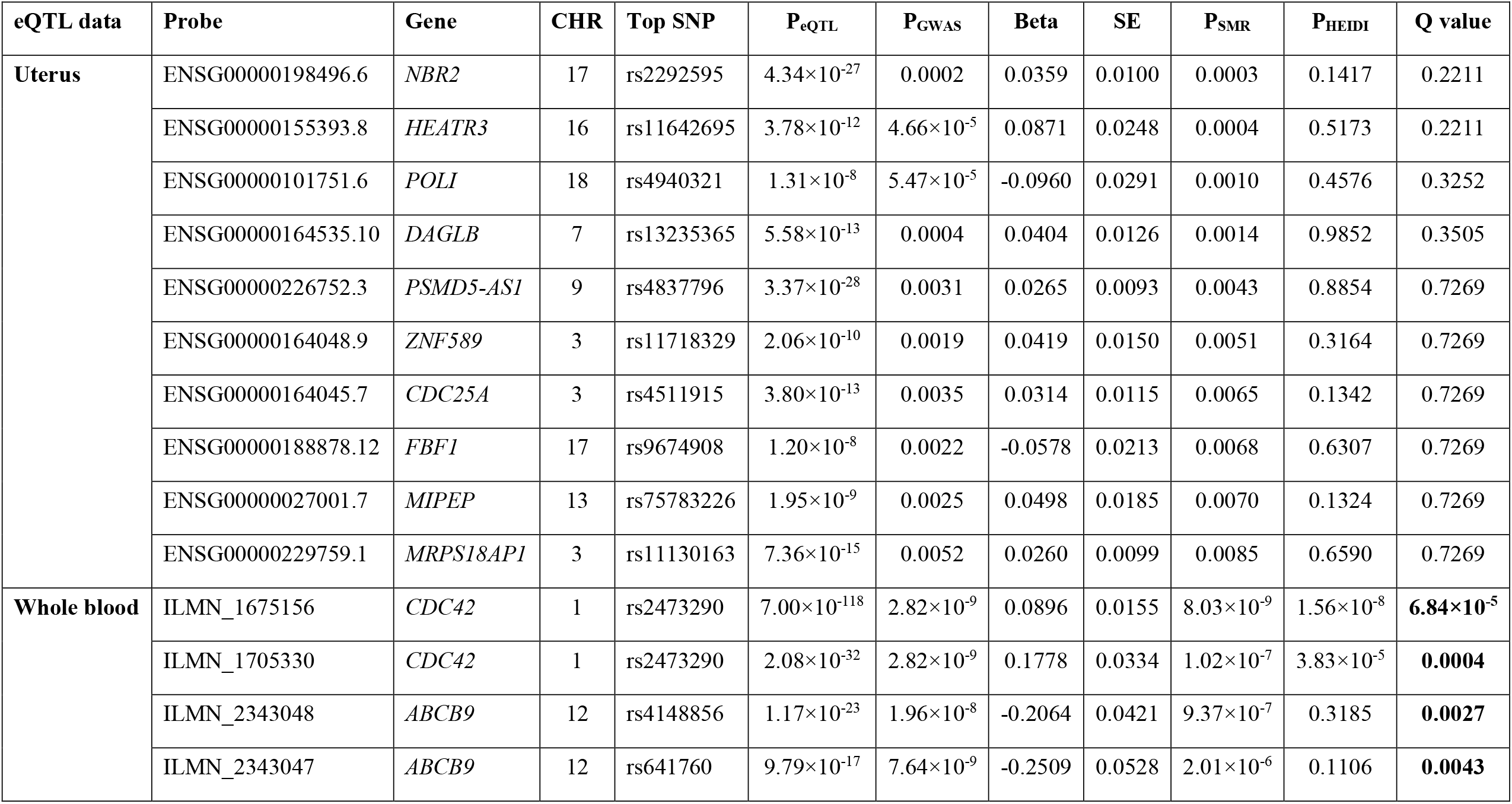
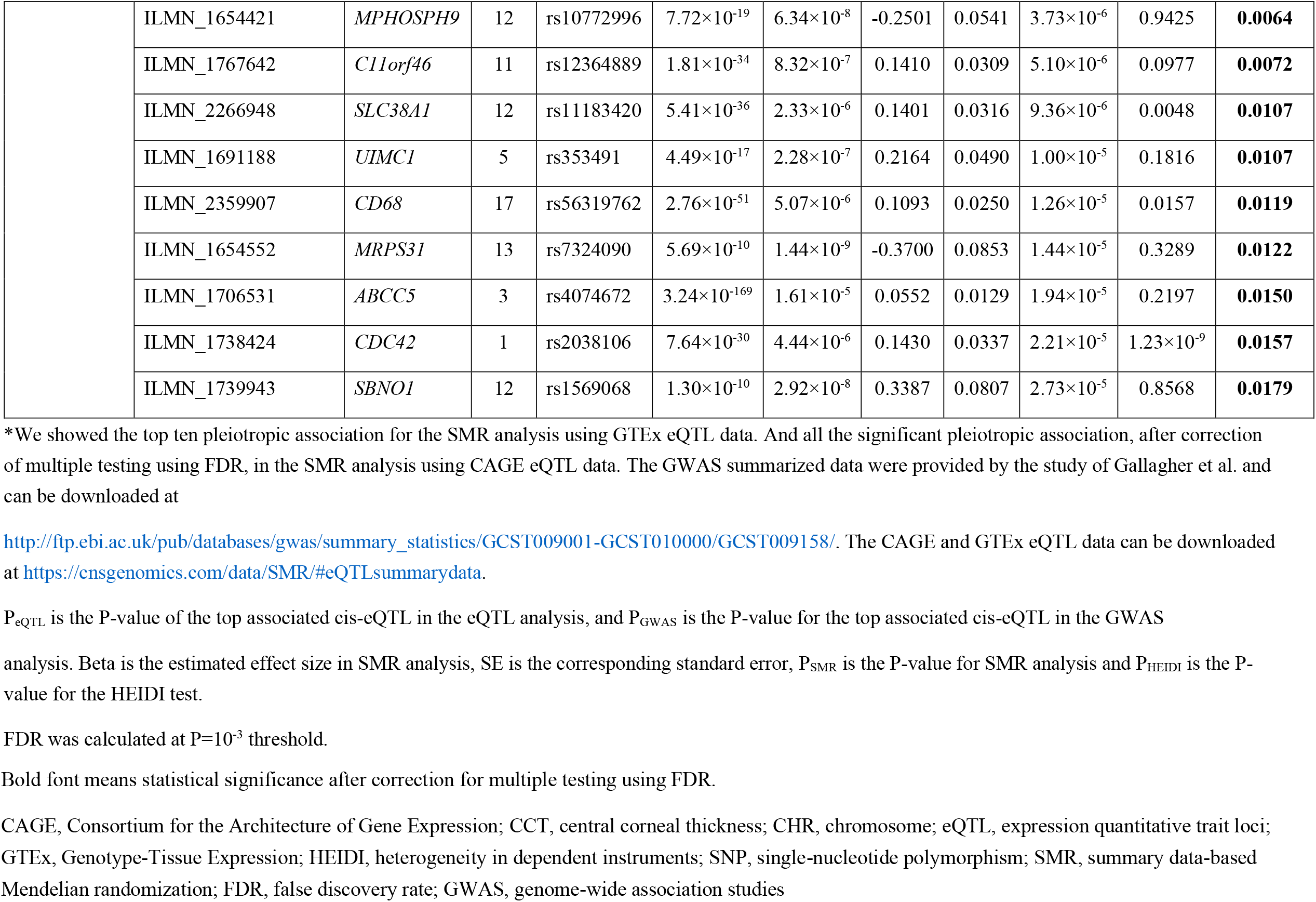
The top hit probes identified in SMR analysis*.

**Figure 2.**
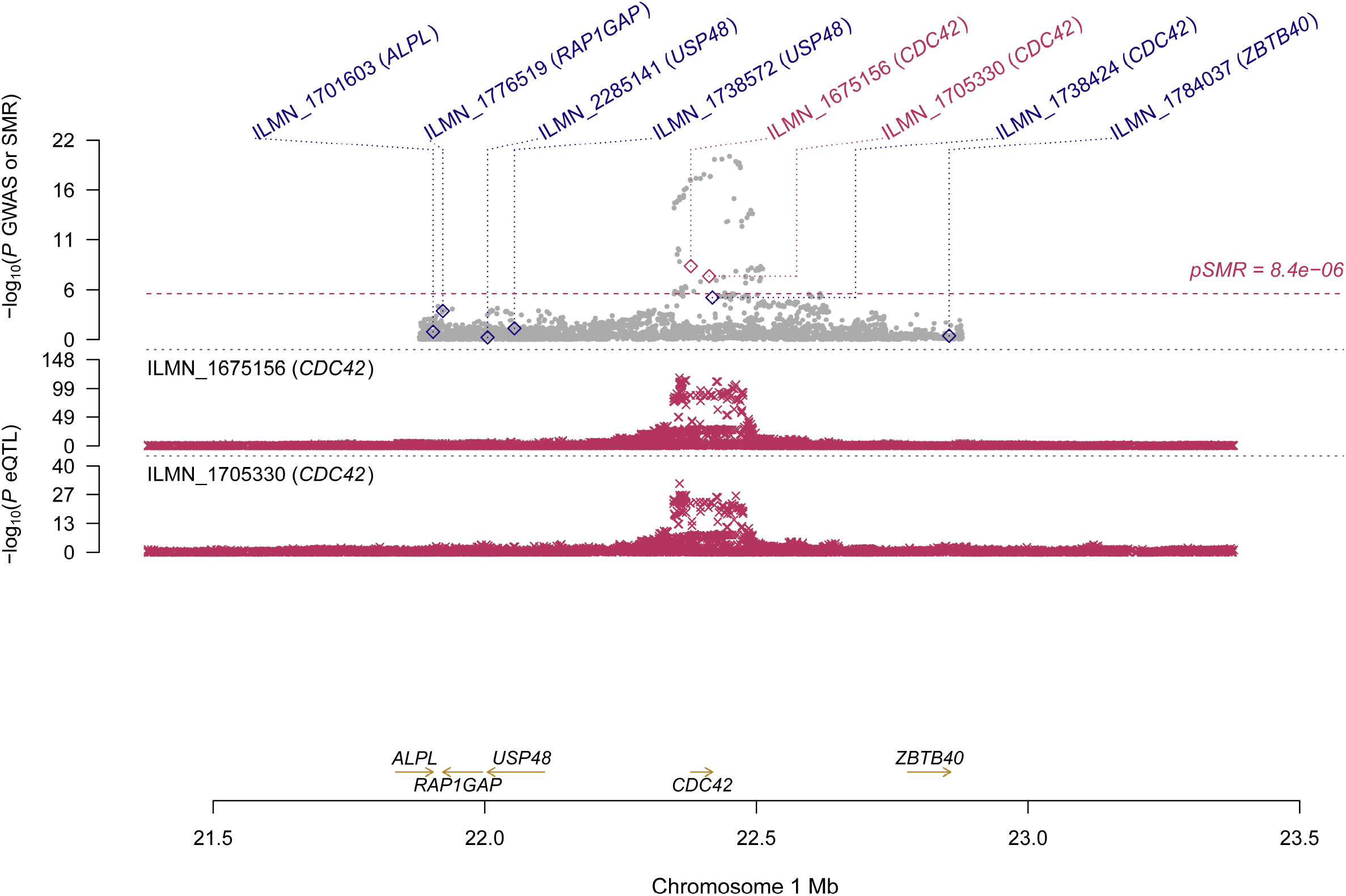
Pleiotropic association of *CDC42* with ULs using CAGE eQTL. Top plot, grey dots represent the −log_10_(P values) for SNPs from the GWAS of ULs, with solid rhombuses indicating that the probes pass HEIDI test. Middle plot, eQTL results. Bottom plot, location of genes tagged by the probes. eQTL, expression quantitative trait loci; GWAS, genome-wide association studies; HEIDI, heterogeneity in dependent instruments; SMR, summary data-based Mendelian randomization; SNP, single nucleotide polymorphism; ULs, uterine leiomyomas

**Figure 3.**
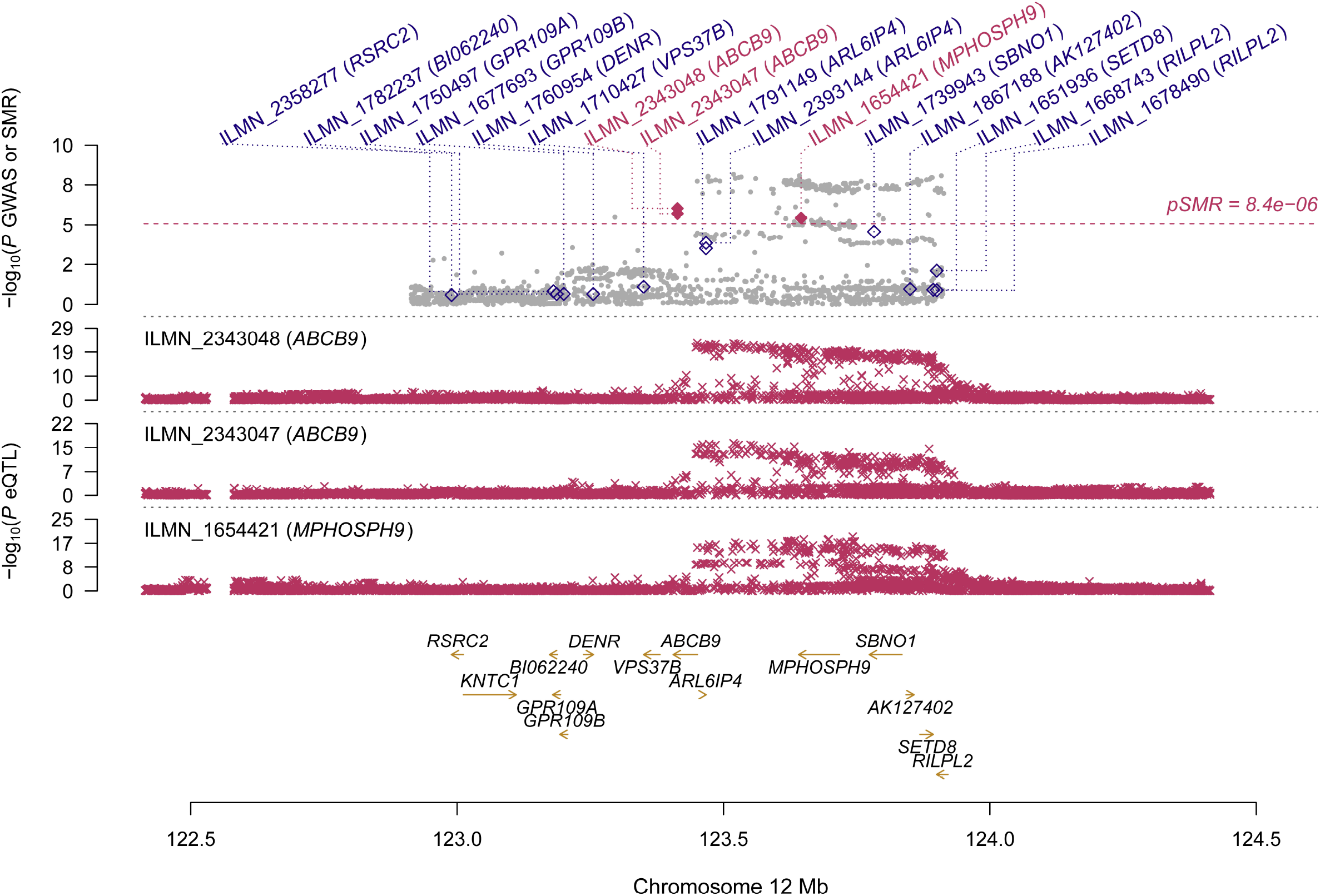
Pleiotropic association of *ABCB9* and *MPHOSPH9* with ULs using CAGE eQTL data. Top plot, grey dots represent the −log_10_(P values) for SNPs from the GWAS of ULs, with solid rhombuses indicating that the probes pass HEIDI test. Middle plot, eQTL results. Bottom plot, location of genes tagged by the probes. eQTL, expression quantitative trait loci; GWAS, genome-wide association studies; HEIDI, heterogeneity in dependent instruments; SMR, summary data-based Mendelian randomization; SNP, single nucleotide polymorphism; ULs, uterine leiomyomas

### Functional mapping and annotation

FUMA analysis identified 106 independent significant SNPs, 33 lead SNPs (**Table S1-S3**), and 24 genomic risk loci (**Figure 4**; **Table S4**). In addition, FUMA identified 137 genes that are potentially involved in the pathogenesis of ULs (**Table S5**). These 137 genes are distributed in 20 genomic risk loci, with four genomic risk loci containing no identified genes (**Figure 4 & Table S5**). Of the 137 identified genes, 7 were also identified by SMR analysis, including *CDC42, SLC38A1, ABCB9, MPHOSPH9, SBNO1, MRPS31* and *CD68*. Expression of the prioritized genes in 30 tissues can be found in **Table S6** and **Figure S1**.

**Figure 4.**
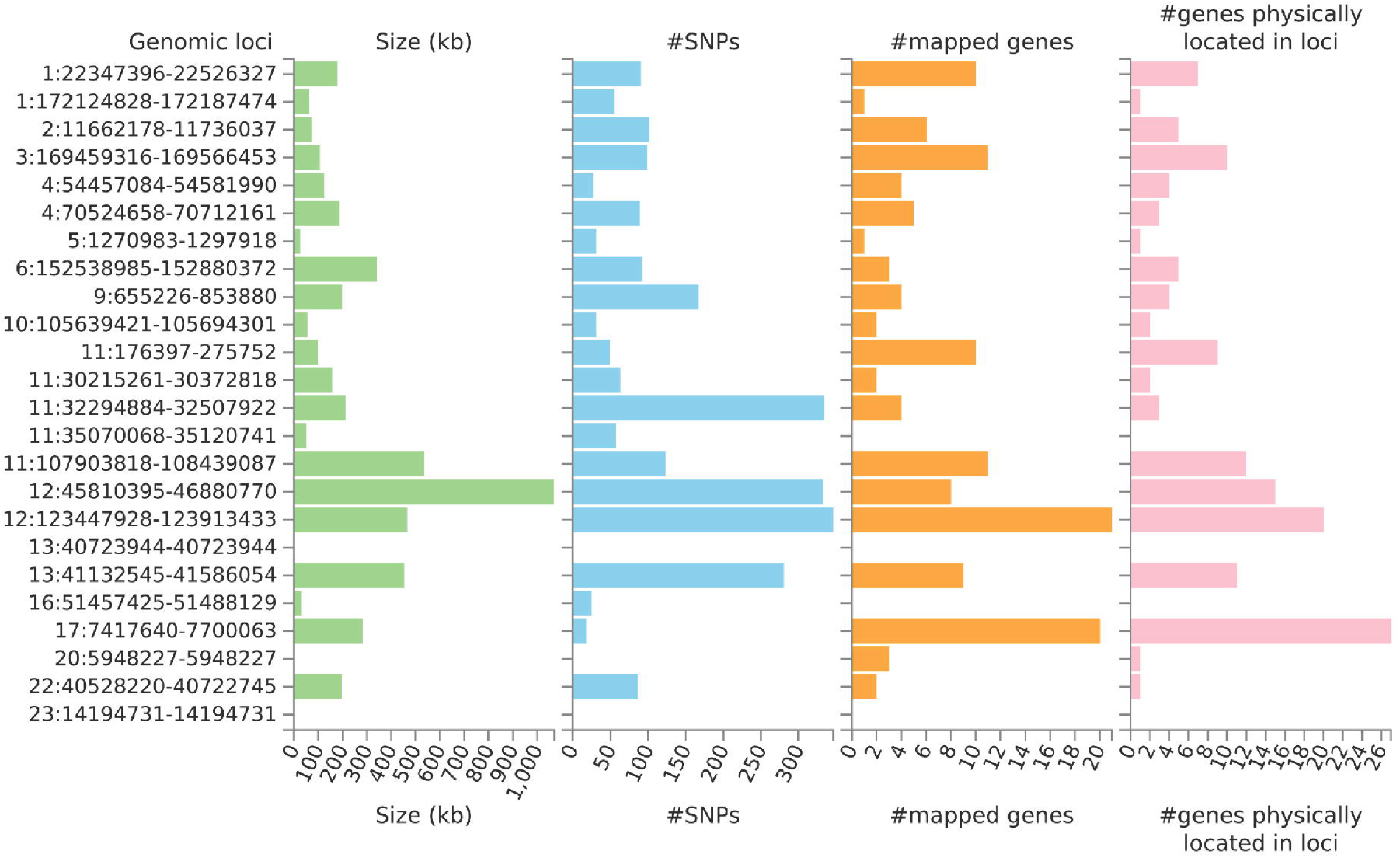
Genetic risk loci identified by FUMA analysis using GWAS data on ULs. Genomic risk loci are displayed in the format of ‘chromosome:start position-end position’ on the Y axis. For each genomic locus, histograms from left to right depict the size, the number of candidate SNPs, the number of mapped genes (using positional mapping and eQTL mapping), and the number of genes known to be located within the genomic locus, respectively. eQTL, expression quantitative trait loci; GWAS, genome-wide association studies; SNP, single nucleotide polymorphism; ULs, uterine leiomyomas

Gene-set enrichment analysis (GSEA) was undertaken to test the possible biological mechanisms of the 137 candidate genes implicated in ULs (**Table S7**). A total of 96 gene sets with an adjusted P < 0.05 were identified. We found enrichment signals related with uterine fibroids (adjusted P=2.00×10^−51^). In addition, we also found enrichment of sex-related signals such as GO_REGULATION_OF_GONAD_DEVELOPMENT (adjusted P=0.023), endometriosis (adjusted P=7.3×10^−4^), sex hormone-binding globulin levels (adjusted P=0.003), and sex hormone levels (adjusted P=0.024; **Table S7**).

## Discussion

In this study, we conducted SMR and FUMA analysis to prioritize SNPs and genes to better understand the genetic mechanisms underlying ULs. We identified multiple genetic variants, genes, genomic risk loci and gene sets that may be involved in the pathogenesis of ULs. These findings provided helpful leads to a better understanding of the pathogenesis of ULs and highlight potential therapeutic targets for the treatment of ULs.

Several probes tagging *CDC42* (cell division control protein 42 homolog) showed significant pleiotropic association with ULs using CAGE eQTL data (**Table 2**) in the SMR analysis using CAGE eQTL data. This gene was also identified by the FUMA analysis. *CDC42* is a member of the Rho family and is implicated in a variety of cellular functions including cell cycle progression, survival, transcription, actin cytoskeleton organization, and membrane trafficking[24]. *CDC42* has been linked to multiple human cancers and is involved in the initiation of many cellular responses during oncogenic processes, such as transition from epithelial to mesenchymal, cell-cycle progression, migration/invasion, tumor growth, angiogenesis, and oncogenic transformation[25, 26]. Several studies reported that *CDC42* might also play an essential role in the pathogenesis of fibroid. For example, genome-wide analysis revealed that the 1p36.12 region, where *CDC42*/*WNT4* is located, was associated with uterine fibroids[13, 14, 16]. Interestingly, the genetic variant rs10917151 in *CDC42*/*WNT4* seems to have ancestry-specific effect on the risk of uterine fibroids. Specifically, the A allele was associated with a reduced risk of uterine fibroids in women of African ancestry (OR=0.84) and an increased risk in women of European ancestry (OR=1.16)[13]. Since rs10917151 has been reported to be involved in hormone-related traits (e.g., endometriosis and endometrial cancer), it probably plays a role in the development of leiomyomas via influencing hormone metabolism[16]. Meanwhile, another study showed that the deregulation of *CDC42* influences fibroblasts activation which is essential in the pathogenesis of ULs[27]. Given the fact that increased cellular proliferation is present in fibroid compared with the adjacent uterine tissue and the function of *CDC42* in influencing cell cycle, further investigation is needed to elucidate the role of *CDC42* in the development of leiomyoma and the potential of this gene as a promising target for the prevention and treatment of ULs.

We also found that two probes tagging *ABCB9* (ATP binding cassette subfamily B member 9) showed significant pleotropic association with ULs in the SMR analysis using CAGE eQTL data. This gene was also identified in the FUMA analysis. *ABCB9* belongs to the superfamily of ATP-binding cassette (ABC) transporters which fulfill diverse physiological functions in different cellular localizations ranging from the plasma membrane to intracellular membranous compartments[28]. *ABCB9*, located on 12q24.31, is an antigen processing-like (TAPL) transporter that has been found to be involved in the development and progression of various malignant tumors, such as ovarian cancer and non-small cell lung cancer[29, 30]. The genetic variant rs2270788 in *ABCB9* was found to be associated with both the risk and tumor size of ULs in African American participants[31]. *ABCB9* was downregulated in women with a high level of progesterone serum (>1.5 ng/ml), compared with women with a lower level of progesterone serum (<1.5 ng/ml)[32]. Since progesterone is a major promoter of leiomyoma development and growth[33], the role of *ABCB9* in fibroids in general, and its function in progesterone-driven growth of leiomyomas in particular, needs further exploration.

The recent GWAS study on ULs also performed two-sample Mendelian randomization analysis[14]. However, the MR analysis was different from our SMR analyses in that their objective was to examine the causality of genetic association between UL and heavy menstrual bleeding (HMB). Their study identified 29 independent loci for ULs, with 27 of them on the autosomal chromosomes while our FUMA analysis identified a total of 24 genomic loci on the autosomal chromosomes. The definition of genomic locus was different between their approach and FUMA: their genomic locus was defined as regions of the genome containing all SNPs in LD (r^2^ > 0.6) with the index SNPs (independent SNPs, i.e., SNPs in low LD (r^2^<0.1) with nearby (≤500 kb) significantly associated SNPs), with any adjacent regions within 250 kb of one another being combined and classified as a single locus. In FUMA, independent significant SNPs (P<5×10^−8^ and independent from each other at r^2^<0.6) were first identified. Then, all known SNPs in LD (r^2^ > 0.6) with one of the independent SNPs were included, using the pre-calculated LD structure based on 1000G. As a results, SNPs that were not originally in the GWAS results could also be included. This may partly explain the difference in the findings.

Our study has limitations. The GWAS analysis was done in participants of European ancestry. As such, our findings might not be generalized to other ethnicities. The number of eligible probes used in SMR analysis was limited, especially in the analysis using GTEx eQTL. As a result, we could not rule out the possibility of missing some important genes that were not tagged in the eQTL data. In addition, the FDR approach to correct for multiple testing resulted in additional possibilities of missing important genes. The HEIDI test was significant for some of the observed associations, implying the existence of horizontal pleiotropy (**Table 2**).

## Conclusions

We identified many genetic variants, genes and genomic loci that are potentially involved in the pathogenesis of ULs. More studies are needed to explore the underlying mechanisms in the etiology of ULs.

## Data Availability

All data generated or analyzed during this study are publicly available as specified in the methods section of this paper. Specifically, the eQTL data can be downloaded at https://cnsgenomics.com/data/SMR/#eQTLsummarydata, and the GWAS summarized data can be downloaded at http://ftp.ebi.ac.uk/pub/databases/gwas/summary_statistics/GCST009001-GCST010000/GCST009158/.

## Declarations

### Ethics approval and consent to participate

This study used only publicly available data. As a result, ethics approval and consent to participate is not needed.

### Consent for publication

Not applicable.

### Availability of data and materials

All data generated or analyzed during this study are publicly available as specified in the methods section of this paper. Specifically, the eQTL data can be downloaded at https://cnsgenomics.com/data/SMR/#eQTLsummarydata, and the GWAS summarized data can be downloaded athttp://ftp.ebi.ac.uk/pub/databases/gwas/summary_statistics/GCST009001-GCST010000/GCST009158/.

### Authors’ contributions

The manuscript was written through contributions of all authors. YD, JY, and LZ designed the study. YD, YZ and JY analyzed data and performed data interpretation. YD, XL and JY wrote the initial draft, and SM, JY and LD contributed to writing subsequent versions of the manuscript. All authors reviewed the study findings and read and approved the final version before submission.

## Acknowledgements

Not applicable

