## Supplementary figures and images for "Exploring potential causal genes for uterine leiomyomas: A summary data-based Mendelian randomization and FUMA analysis"

### FigureS1_mod_030222.tif

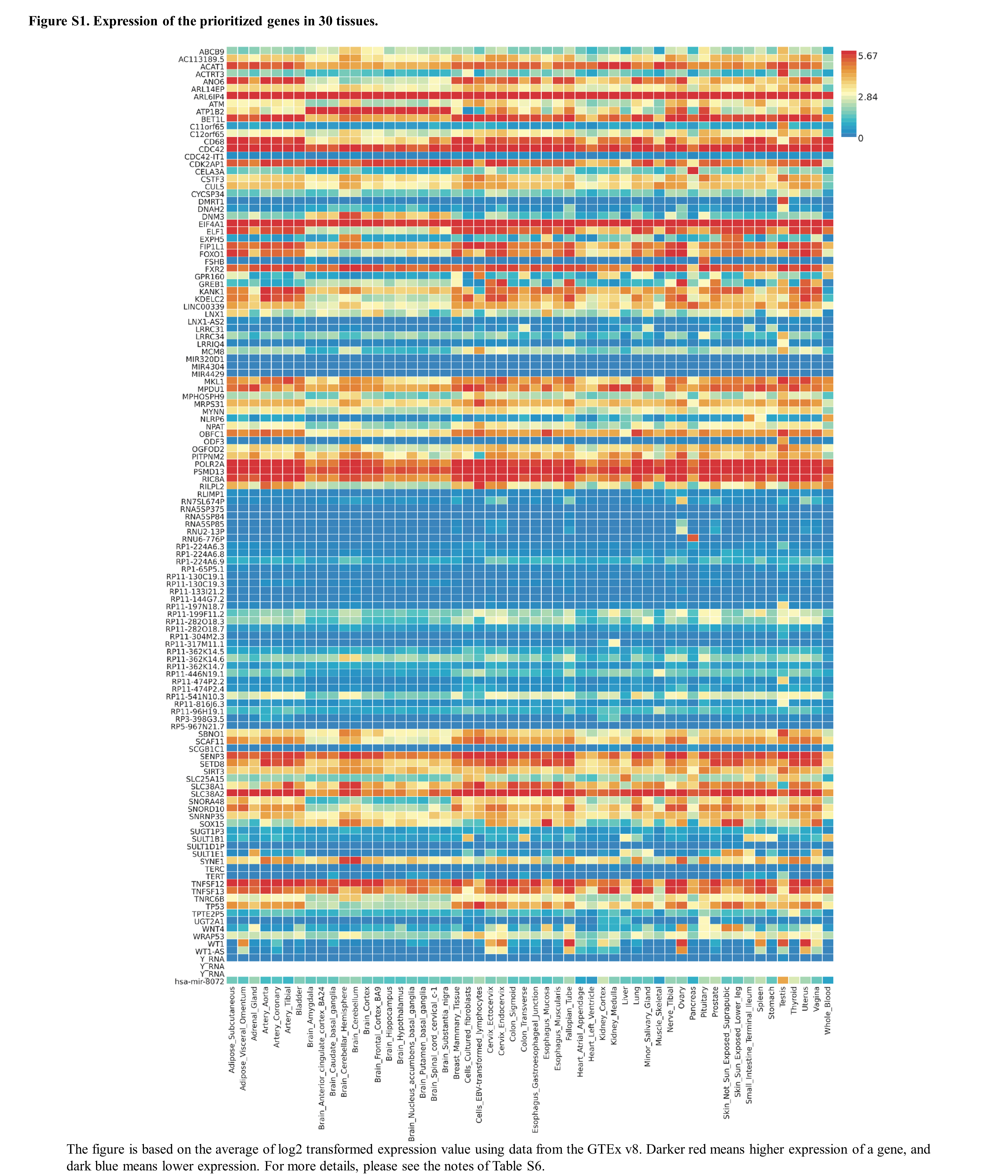
